# Coronary Artery Bypass Grafting and Atrial Fibrillation: Causal Insights from Mendelian Randomization and Retrospective Cohort Analysis

**DOI:** 10.1101/2025.03.20.25324342

**Authors:** Xu Jiacheng, Li Li, Wang Haizhen, Wang Jiamao, Shan Zhonggui

**Affiliations:** Department of Cardiac Surgery, First Affiliated Hospital of Xiamen University, School of Medicine, Xiamen University, Xiamen, China; Department of Anesthesiology, First Affiliated Hospital of Xiamen University, School of Medicine, Xiamen University, Xiamen, China; Teaching Hospital of Fujian Medical University, Department of Cardiovascular Surgery, The First Affiliated Hospital of Xiamen University, Xiamen, China

**Keywords:** Coronary Artery Bypass Grafting, Atrial fibrillation, Mendelian randomization, Medical Information Mart for Intensive Care IV

## Abstract

**Background:** This study aimed to investigate the causal relationship between coronary artery bypass grafting (CABG) and atrial fibrillation (AF) using Mendelian Randomization (MR) and a retrospective cohort from the Medical Information Mart for Intensive Care IV (MIMIC-IV) database.

**Methods:** We performed two-sample MR analysis using GWAS summary statistics (UK Biobank and EBI) to identify genetic instruments for CABG, followed by sensitivity analyses (MR-Egger, weighted median) to validate causality. Concurrently, 1, 835 ICU patients from MIMIC-IV were analyzed via multivariable logistic regression to assess CABG-AF association, stratified by age, hemodynamic, and coagulation profiles.

**Results:** MR analysis demonstrated a robust causal effect of CABG on AF (IVW OR=1.8, P=2.33×10⁻⁹), corroborated by cohort data showing doubled AF risk post-CABG (OR=2.1, 95% CI:1.4–3.1, P<0.001). Subgroups with autonomic instability (low heart/respiratory rates) or coagulopathy (INR>2.5) exhibited heightened susceptibility.

**Conclusion:** CABG independently elevates AF risk via autonomic, inflammatory, and hemostatic pathways, necessitating tailored perioperative monitoring and prophylactic interventions.

## Introduction

CABG is a common surgical procedure performed to restore blood flow to the heart in patients with coronary artery disease. AF is the most sustained common arrhythmia, and its incidence and prevalence are increasing in globally(1).The elegant studies had reported that incidence of AF post CABG ranges from 20% to 50% and associated with higher adverse effect such as stroke, prolonged hospitalization, and increased mortality (2, 3). Despite the high incidence of atrial fibrillation (AF) after coronary artery bypass grafting (CABG), the causal mechanisms remain unclear, particularly for new-onset AF (NOAF) (4).

Mendelian Randomization(MR) is a method used in epidemiology and genetic research to investigate causal relationships between risk factors and outcomes(5).In this approach, genetic variants that are associated with the exposure but not directly associated with the outcome are used as instrumental variables.These genetic variants are randomly assigned at conception and are not influenced by confounding factors or reverse causality.

Medical Information Mart for Intensive Care IV(MIMIC-IV) is a publicly available critical care database that contains de-identified health data from patients admitted to intensive care units (ICUs).The database has been instrumental in advancing research in critical care medicine, machine learning applications in healthcare, and clinical informatics.By leveraging the high-quality, large sample size, and nationally representative nature of the data in MIMIC-IV, robust analyses can be assessed the relationship between CABG and AF.

In this study, we combined a large-scale retrospective cohort analysis using the MIMIC-IV database with a two-sample Mendelian Randomization approach to comprehensively evaluate the causal relationship between CABG and AF. This dual-method approach allows for robust assessment of the association while addressing potential confounding and biases inherent in observational studies. By integrating genetic and clinical data, we aim to provide deeper insights into the mechanisms underlying the development of AF following CABG and inform strategies for risk mitigation and patient management.

## Methods

### Mendelian Randomization analysis

#### Overall study design and data sources

The study employed a retrospective cohort design to investigate the causal relationship between CABG and atrial fibrillation using MR analysis.Research Ethics Committee approved and waived the requirement due to the absence of personal data. Access to open and easily obtainable GWAS data is indeed advantageous for this research reproducibility and accuracy. All details of datasets employed in our study are displayed in Table 1.

**Table 1.** Characteristics of enrolled GWASs in CABG and AF.

| Characteristics | CABG | AF |
| --- | --- | --- |
| No.of controls | 60,620 | 970,216 |
| No.of cases | 3,511 | 60,620 |
| Sample size | 462,933 | 1,030,836 |
| Year of publication | 2018 | 2018 |
| Number of SNPs | 9,851,867 | 33,519,037 |
| Study population | European | European |

The CABG-related GWAS data which characterized by cooperation code(coronary arterial bypass graft) were obtained from the UK Biobank.The UK Biobank is indeed a large-scale and detailed cohort study that has been instrumental in advancing biomedical research(6). The Summary-level GWAS data for AF were gained from European Bioinformatics Institute with up to 1030836 participants(60620 cases with AF and 970216 cases without AF)(Table 1 shown). **Instrument variable (IVs) selection.**

According to our MR study design (Figure 1 shown), we selected a set of screening criteria in order to identify applicable IVs associated with CABG and AF. Well-powered instrumental variables (IVs) should ideally fulfill the three fundamental assumptions as following:(i) IVs should be strongly associated with the exposure of interest, we adopted candidate SNPs strongly associated with CABG as IVs from GWAS data in this study(p<5×10^-8^);(ii)IVs should be independent of confounding factors that could bias the causal estimates, we seted strict conditions(r^2^>0.001 and physical distance < 10, 000 kb) to avoid bias caused by linkage disequilibrium among SNPs;(iii) IVs should only affect the outcome through their effect on the exposure and should not have any direct effect on the outcome that is independent of the exposure, we removed SNPs (F<10) to mitigate bias associated with weak IVs.

**Figure I.**
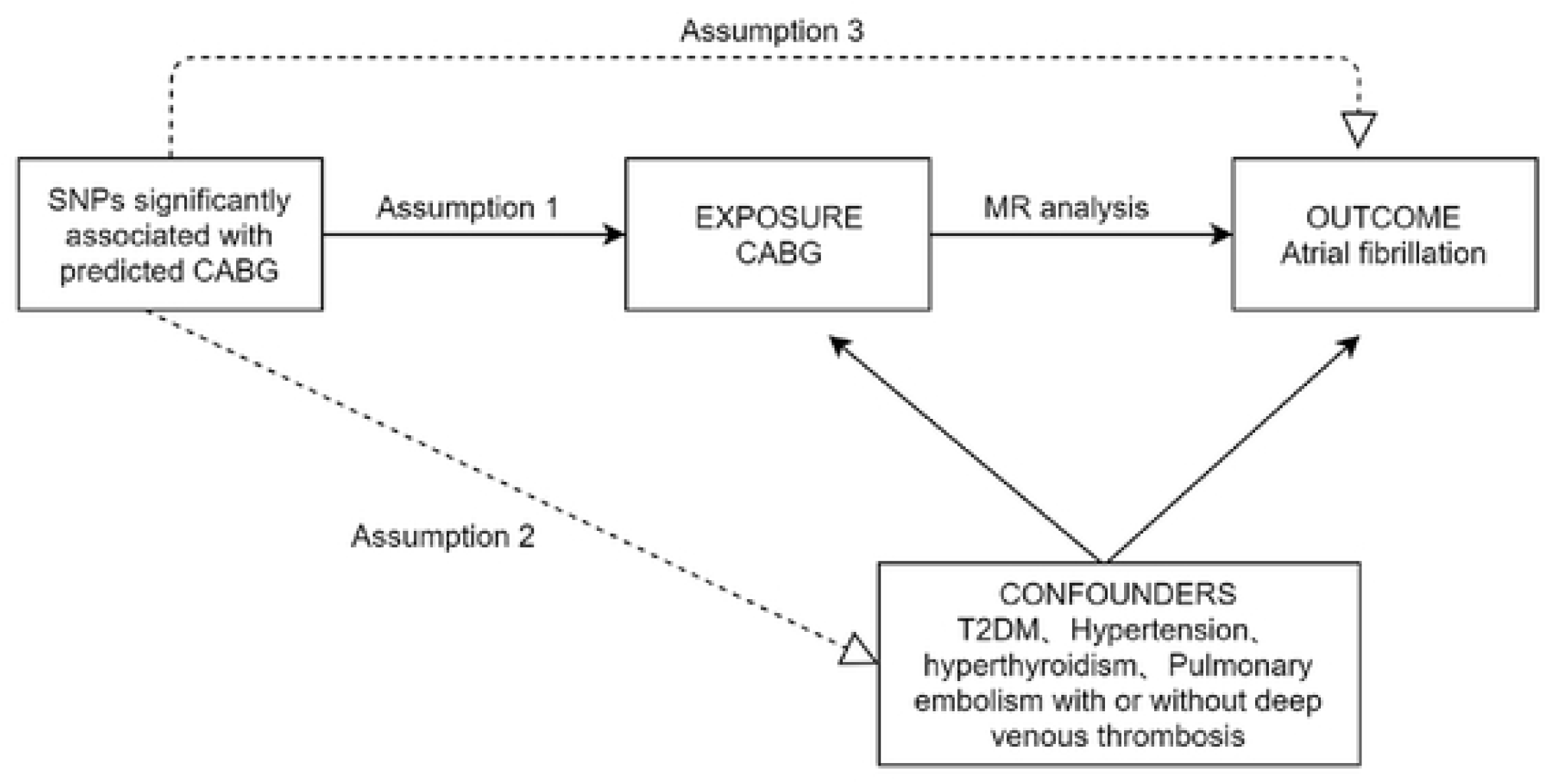
Three imperative assumptions of Mendelian randomization approach.SNP, singlenucleotide polymorphism;CABG, cardiac arterial bypass graft;T2DM, type 2 diabetes mellitus.

### MIMIC-IV retrospective study Data section and flowchart

The data used in the current analysis are publicly available through the MIMIC-IV v3 database(https://physionet.org/content/mimiciv/3.0/). The process of selecting patients with coronary atherosclerosis admitted to the ICU for the first time. Exclusion criteria included patients who did not undergo surgical intervention with CABG or PCI and combined hyperthyroidism or mitral valve disease (Figure 3 shown).In order to obey with the relevant rules, the author Li Li(record number:13369960) joined and completed online the course on protecting human research participants proposed by the US National Institutes of Health’s, and therefore obtained permission to access the dataset. The Structured Query Language (SELECT) of Navicat Premium (Version 16) was employed to extracted baseline characteristics data, such as age, gender, ethnicity, laboratory indicators, and comorbidities.

### Clinic definitions

The risk of postoperative atrial fibrillation was the main outcome of this study. Atrial fibrillation was defined by the ICD-10 codes.Secondary outcome was 28-day survival rate after CABG or PCI.

### Statistical analysis

The study adopted a recently elegant two-sample MR analysis of three different approach, including inverse variance weighted(IVW), MR-Egger and weighted median regression, calculate the causal correction between CABG and AF. Firstly, the IVW estimated the causal effect due to its highly statistical power through pooling the Wald ratio estimates of each SNP on the outcome. Subsequently, MR-Egger and weighted median regression were employed to ensure stability and reliability of the MR results(7).MR-Egger intercept test(p<0.05 was considered to be pleiotropy), MR-Pleiotropy Residual Sum and Outlier methods(MR-PRESSO) and funnel plot were further applied to detect potential pleiotropy of MR estimates. It’s worth noting that MR-PRESSO causes regression in the estimates of SNP-exposure on the estimates of SNP-outcome, aiming to detect outlier SNPs and yield calibrated causality(8).Furthermore, Cochrane’s Q test was used to assess heterogeneity among SNPs based on IVW and MR-Egger analysis and p<0.05 were presented for significance of heterogeneity(9).Lastly, leave-one-out(LOO) analysis was used to evaluate whether any single SNP determined causality and the Steiger directionality test was applied to determine the direction of causal effect(p<0.05 was of significance)(10).

To eliminate the core assumption that confounding factors affect MR causal results, we screened for SNPs(level of value was 1X10^-5^) associated with risk factors for atrial fibrillation, including BMI, type 2 diabetes mellitus, hyperthyroidism or thyrotoxicosis, hypertension and Pulmonary embolism with or without deep venous thrombosis(11).We will eliminate and recompute MR analysis to verify results consistency when any IVs associated with potential confounder. All statistical analyses were performed using R software (version 4.2.1) with the “Two-Sample MR” and “MRPRESSO” R packages.

In MIMIC-IV database, the distribution of baseline data was shown as different result groups for patients involved in this study(Table 3 shown). The results are presented as Mean ± SD/N(%). For continuous variables, differences between groups were assessed using the Kruskal-Wallis rank sum test. For categorical variables with theoretical counts less than 10, the Fisher’s exact test was employed. Assessment of the independent association between CABG and the risk of atrial fibrillation was conducted using a multivariate logistic regression analysis. The CABG exposure factor was input as a categorical variable (quartile) and a continuous variable (a risk ratio (OR) was calculated for each additional unit). We further conducted a hierarchical analysis based on age, gender, SBP, resp rate, heart rate, SpO2, WBC, monocytes, neutrophils, lactate, glucose, fibrinogen, INR, PT, PTT, CRRT, ventilator, so as to determine the robustness of the CABG in predicting the risk of atrial fibrillation.Furthermore, three models were used in the regression analysis. Adjustments were made to the multivariable models, as follows: No adjustments were made to Model 1; adjustments were made for Model 2 according to age, gender, race, BMI; Model 3 was adjusted according to Model 2 by adding SBP, MBP, resp rate, heart rate, SpO2, WBC, monocytes, neutrophils, lactate, glucose, fibrinogen, INR, PT, PTT, CRRT, ventilator.

## Results

### Results of MR analysis

In the present study, after strict primary selection process, we chosen SNPs association with Coronary Artery Bypass Grafting(CABG)(P<5×10^-8^) and available in the NOAF dataset. The value of F-statistic for all chosen SNPs were above 10, indicating no weak IVs for this MR analysis.

The two-sample MR analysis methods were adopted to explore the genetically predicted associations between CABG and the episodes of AF(Figure 2).We found robust evidence of causal estimate of CABG on AF through IVW and CABG was positively associated with AF(IVW:P=2.33×10^-9^).Similarly, MR-Egger regression(P=0.04) and weighted median method(P=3.64×10^-6^) had shown the consistent estimate in this study.Furthermore, we used other approachs(Simple mode(P=0.014) and Weighted mode methods(P=0.007)) to detect positive causality and strengthen the reliability of our analysis.

**Figure 2.**
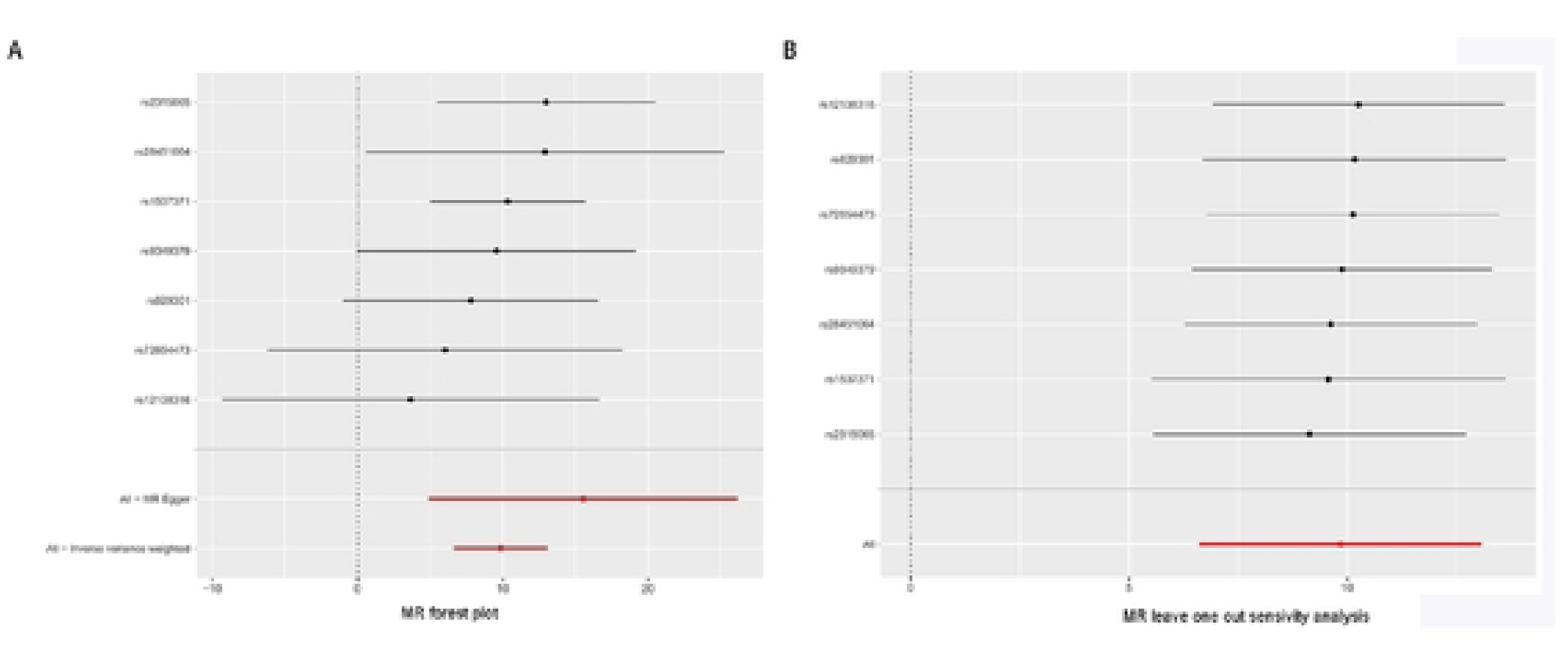
A. Forest plot to visualize causal effect of each single SNP on NOAF risk.B.Leave­ one-out plot to visualize causal effect of CABG on NOAF when leaving one SNP out.

To further explore causality, we conducted a directional test by MR Steiger. The result had shown that there was no reverse causality between CABG and AF (P<0.05).

The results of Cochrans’ Q test displayed no heterogeneity among all SNPs(IVW:Q=2.39, P=0.88;MR-Egger:Q=1.19, P=0.95).The value of MR-Egger intercept test showed no evidence of possible horizontal pleiotropy(intercept=1.13×10^-2^, P=0.32). In addition, no potential pleiotropy was found by using the MR-PRESSO global test (P = 0.899). Therefore, there were no directional pleiotropic effects of CABG on AF.The funnel plot and leave-one-out indicted a steady estimate of IVW results. Power analysis provided sufficient power (100%) to infer the impact of CABG on AF in massive sample size (1030836) and the threshold of 0.05. To further eliminated potential confounders factors connecting AF, we adopted multivariable MR analysis to adjust the effects on this indicators. The values of MVMR analysis showed that CABG still significantly corrected with the episodes of AF(Table 2 shown).

**Table 2.** The multivariable Mendelian randomization (MVMR) analysis to investigate the causal relationship between CABG and AF.

| Key Confounding factors | P value | Odds Ratios(OR) |
| --- | --- | --- |
| T2DM | 0.17 | 0.98(0.95-1.01) |
| Hypertension | 0.56 | 0.15(0-93.07) |
| Hyperthyroidism | 0.70 | 1.73(0.10-28.54) |
| PE(DVT) | 0.10 | 27.73(0.56-1380.10) |

T2DM: type 2 diabetes mellitus, PE(DVT): Pulmonary embolism with or without deep venous thrombosis.

### Results of MIMIC-IV cohort study

A total of 1835 patients were eventually involved and were divided into two groups according to exposure factors in this study(Figure 3 shown).1464 patients were enrolled in CABG group, the average age was 69.0 ± 10.6 years old, and 1097 (74.9%) were female. The incidence of atrial fibrillation in CABG group was 39.8%(n=582)(Table 3 shown). The figure 4 presents that the probability of survival is higher for patients who underwent CABG compared to those who underwent PCI, with a statistically significant difference indicated by a p-value of less than 0.01 (P<0.01).

**Figure 3.**
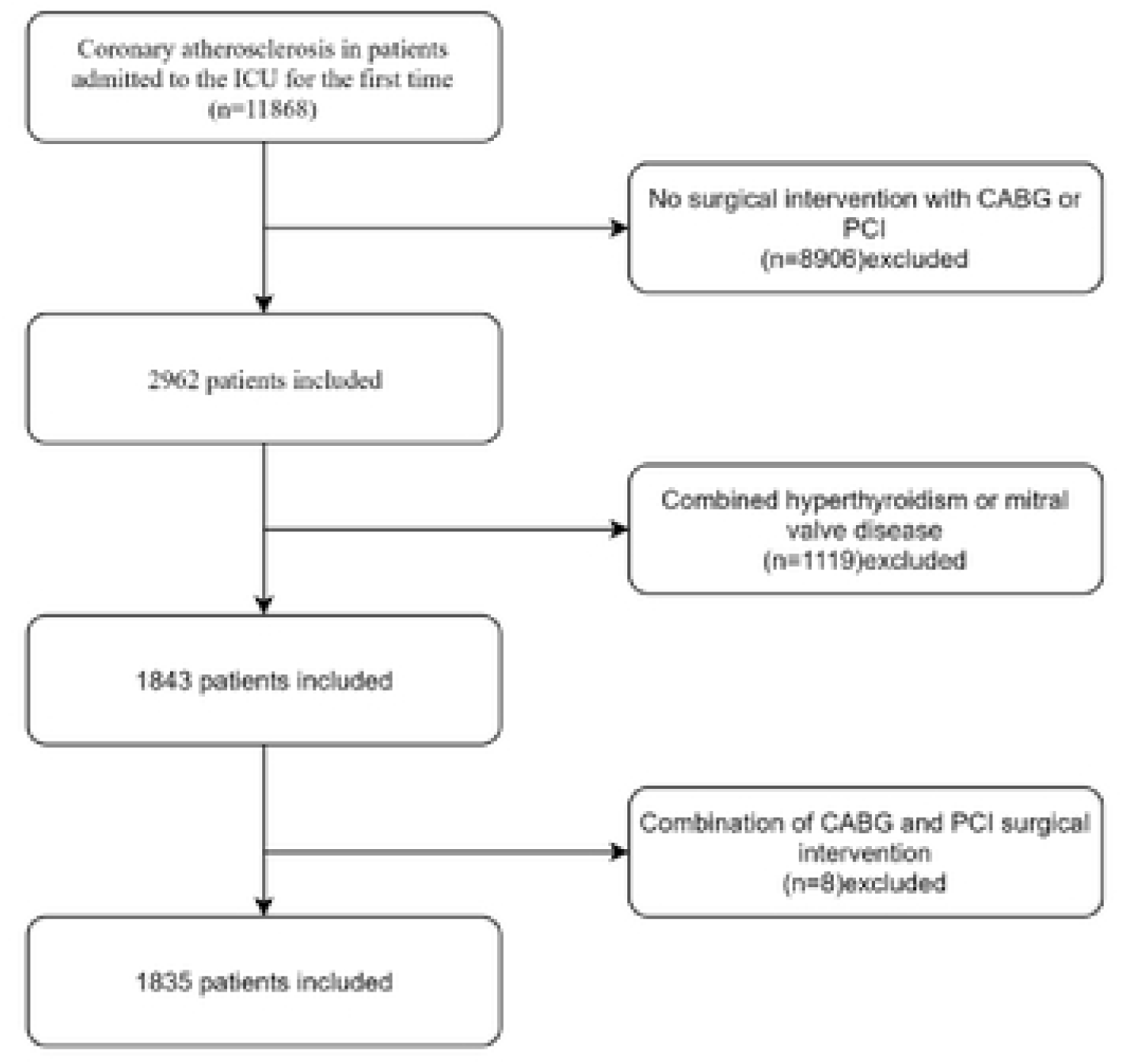
A flowchart detailing the selection process of patients with coronary atherosclerosis disease.

**Figure 4.**
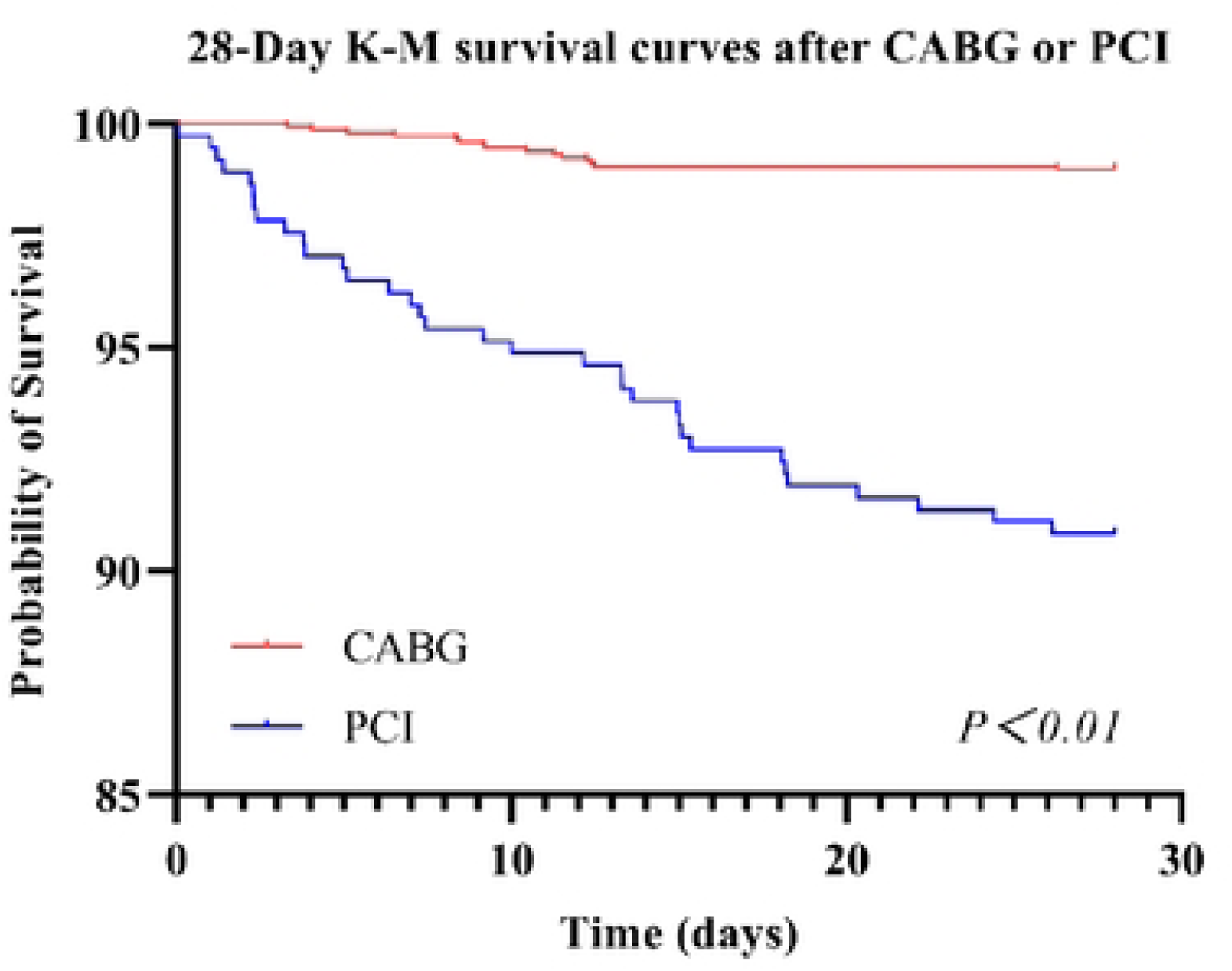
The figure presents the 28-day Kaplan-Meier **(K-M)** survival curves for patients who underwent either CABG or PCI.

**Table 3.** Baseline characteristics of participants.

| Variables | PCI (n=371) | CABG (n=1464) | Standardize Diff. | P-Value |
| --- | --- | --- | --- | --- |
| Age, years | 71.5 ± 12.5 | 69.0 ± 10.6 | 0.2 (0.1, 0.3) | <0.001 |
| Gender, sex |  |  | 0.3 (0.1, 0.4) | <0.001 |
| Female, n (%) | 235 (63.3%) | 1097 (74.9%) |  |  |
| Male, n (%) | 136 (36.7%) | 367 (25.1%) |  |  |
| Race |  |  | 0.1 (0.0, 0.3) | 0.345 |
| Asian, n (%) | 8 (2.2%) | 23 (1.6%) |  |  |
| Black, n (%) | 18 (4.9%) | 49 (3.3%) |  |  |
| White, n (%) | 261 (70.4%) | 1046 (71.4%) |  |  |
| Hispanic, n (%) | 15 (4.0%) | 49 (3.3%) |  |  |
| Unknown, n (%) | 62 (16.7%) | 244 (16.7%) |  |  |
| Other, n (%) | 7 (1.9%) | 53 (3.6%) |  |  |
| BMI, kg/m<sup>2</sup> | 28.9 ± 6.3 | 29.3 ± 5.8 | 0.1 (-0.0, 0.2) | 0.141 |
| SBP (mmHg) | 127.6 ± 24.5 | 114.7 ± 18.3 | 0.6 (0.5, 0.7) | <0.001 |
| MBP (mmHg) | 83.6 ± 16.9 | 78.1 ± 13.2 | 0.4 (0.2, 0.5) | <0.001 |
| Resp Rate, bpm | 18.3 ± 5.1 | 15.1 ± 3.7 | 0.7 (0.6, 0.8) | <0.001 |
| Heart Rate, bpm | 80.9 ± 17.8 | 80.0 ± 10.5 | 0.1 (-0.1, 0.2) | 0.229 |
| WBCs (10<sup>9</sup>/L) | 10.7 ± 5.4 | 11.3 ± 5.0 | 0.1 (0.0, 0.2) | 0.006 |
| Monocytes (10<sup>9</sup>/L) | 3.8 ± 2.1 | 3.3 ± 1.8 | 0.3 (0.2, 0.4) | <0.001 |
| Neutrophils (10<sup>9</sup>/L) | 77.7 ± 12.6 | 77.2 ± 10.5 | 0.0 (-0.1, 0.2) | 0.037 |
| Lactate (mmol/L) | 1.9 ± 1.5 | 1.3 ± 0.6 | 0.5 (0.4, 0.6) | <0.001 |
| Glucose (mg/dL) | 168.3 ± 84.9 | 138.9 ± 40.8 | 0.4 (0.3, 0.6) | <0.001 |
| Fibrinogen (mg/dL) | 237.9 ± 91.3 | 232.1 ± 81.5 | 0.1 (-0.0, 0.2) | 0.408 |
| INR | 1.3 ± 0.6 | 1.3 ± 0.3 | 0.0 (-0.1, 0.1) | <0.001 |
| PT (s) | 14.6 ± 5.8 | 14.8 ± 2.7 | 0.0 (-0.1, 0.1) | <0.001 |
| PTT (s) | 48.6 ± 31.7 | 39.1 ± 20.4 | 0.4 (0.2, 0.5) | <0.001 |
| Ventilator (hours) | 69.1 ± 96.3 | 50.7 ± 79.1 | 0.2 (0.1, 0.3) | 0.927 |
| LOS ICU (days) | 3.1 ± 3.7 | 2.7 ± 3.7 | 0.1 (-0.0, 0.2) | 0.516 |
| LOS Hospital (days) | 8.0 ± 7.0 | 10.0 ± 5.6 | 0.3 (0.2, 0.4) | <0.001 |
| SpO<sub>2</sub> (%) | 96.7 ± 3.5 | 99.0 ± 2.0 | 0.8 (0.7, 0.9) | <0.001 |
| CRRT |  |  | 0.2 (0.1, 0.3) | <0.001 |
| No, n (%) | 356 (96.0%) | 1445 (98.7%) |  |  |
| Yes, n (%) | 15 (4.0%) | 19 (1.3%) |  |  |
| Atrial Fibrillation |  |  | 0.3 (0.2, 0.5) | <0.001 |
| No, n (%) | 282 (76.0%) | 882 (60.2%) |  |  |
| Yes, n (%) | 89 (24.0%) | 582 (39.8%) |  |  |
| 28-Day Mortality |  |  | 0.4 (0.3, 0.5) | <0.001 |
| No, n (%) | 337 (90.8%) | 1449 (99.0%) |  |  |
| Yes, n (%) | 34 (9.2%) | 15 (1.0%) |  |  |

The results are presented as Mean ± SD / N (%). For continuous variables, differences between groups were assessed using the Kruskal-Wallis rank sum test. For categorical variables with theoretical counts less than 10, the Fisher's exact test was employed.
Abbreviations: PCI, percutaneous coronary intervention; CABG, coronary artery bypass grafting; BMI, body mass index; WBC, white blood cell; SpO<sub>2</sub>, peripheral oxygen saturation; SBP, systolic blood pressure; MAP, mean artery pressure; LOS ICU, length of stay in ICU; LOS hospital, length of stay at the hospital; CRRT, continuous renal replacement therapy.

The overall analysis demonstrated a statistically significant association between CABG and the development of AF, with an odds ratio (OR) of 2.1 (95% CI: 1.4–3.1, P < 0.001), indicating that patients undergoing CABG had more than double the risk of AF compared to those who did not(Table 4 shown). In the subgroup analysis based on age, the effect of CABG on AF varied across tertiles (Table 5 shown). In the low tertile, the association was not statistically significant (OR: 1.8, 95% CI: 1.0–3.3, P = 0.059). However, in the middle (OR: 2.6, 95% CI: 1.6–4.3, P < 0.001) and high tertiles (OR: 2.7, 95% CI: 1.8–3.9, P < 0.001), the association was significant. Although there was no statistically significant interaction between age and the effect of CABG on AF (P for interaction = 0.109), there was a trend suggesting a higher risk with increasing age. Both female (OR: 2.0, 95% CI: 1.4–2.7, P < 0.001) and male (OR: 2.4, 95% CI: 1.5–3.8, P < 0.001) patients exhibited a significant association between CABG and AF. The risk appeared slightly higher in males, though the interaction between gender and CABG was not statistically significant (P for interaction = 0.281). The analysis across BMI tertiles revealed significant associations in all groups: low (OR: 1.7, 95% CI: 1.1–2.6, P = 0.010), middle (OR: 2.4, 95% CI: 1.5–3.8, P < 0.001), and high (OR: 2.3, 95% CI: 1.4–3.7, P < 0.001). However, no significant interaction was observed between BMI and the CABG–AF relationship (P for interaction = 0.375). In the SBP subgroup, the low tertile showed no significant association between CABG and AF (OR: 1.3, 95% CI: 0.8–2.1, P = 0.343). Conversely, both the middle (OR: 2.1, 95% CI: 1.3–3.5, P = 0.004) and high tertiles (OR: 2.5, 95% CI: 1.7–3.7, P < 0.001) demonstrated significant associations, with a trend of increasing risk at higher SBP levels. Although the interaction term approached significance, it did not reach statistical significance (P for interaction = 0.081). A notable finding was observed in the respiratory rate tertiles. Both the low (OR: 3.8, 95% CI: 1.9–7.7, P < 0.001) and middle tertiles (OR: 3.5, 95% CI: 1.7–7.2, P < 0.001) showed stronger associations compared to the high tertile (OR: 1.7, 95% CI: 1.2–2.3, P = 0.002). The interaction between respiratory rate and the CABG– AF relationship was statistically significant (P for interaction = 0.033), indicating that respiratory rate may modify the effect of CABG on AF development. Heart rate also demonstrated a significant interaction (P for interaction = 0.031). The association was strongest in the low heart rate tertile (OR: 2.6, 95% CI: 1.7–4.0, P < 0.001) and decreased in the middle (OR: 2.3, 95% CI: 1.2–4.2, P = 0.010) and high tertiles (OR: 1.6, 95% CI: 1.1–2.3, P = 0.027).

**Table 4.**
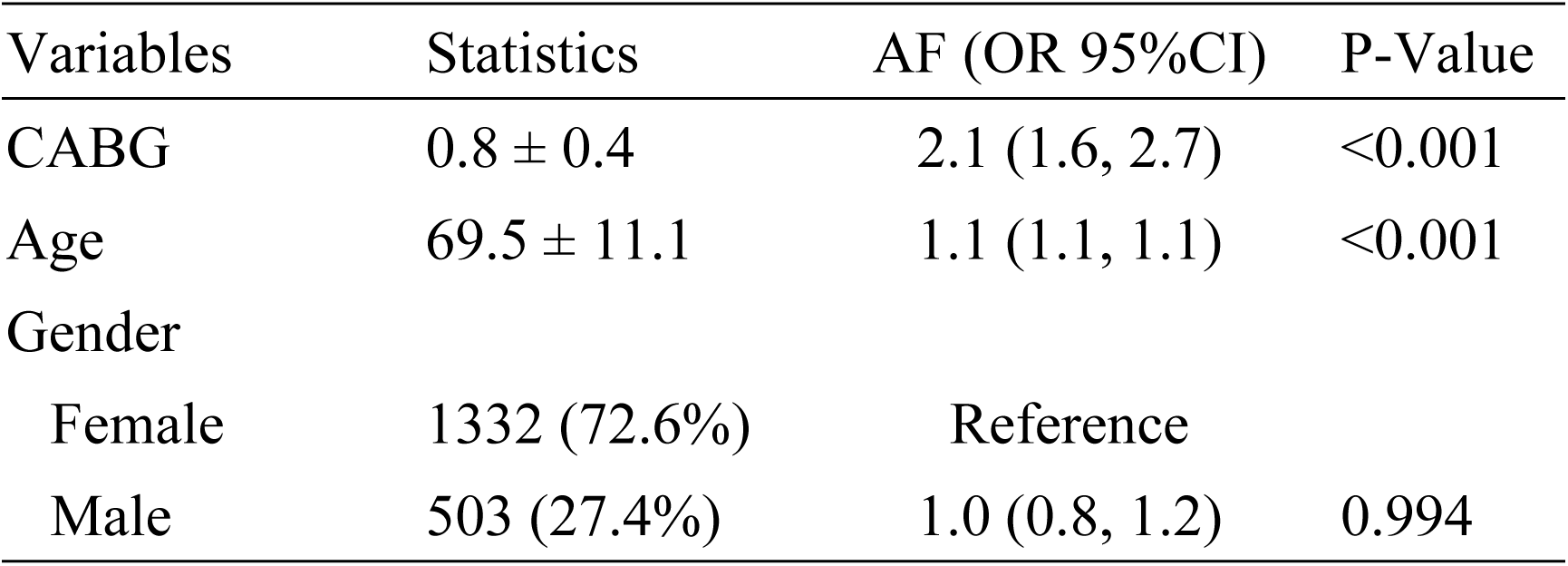
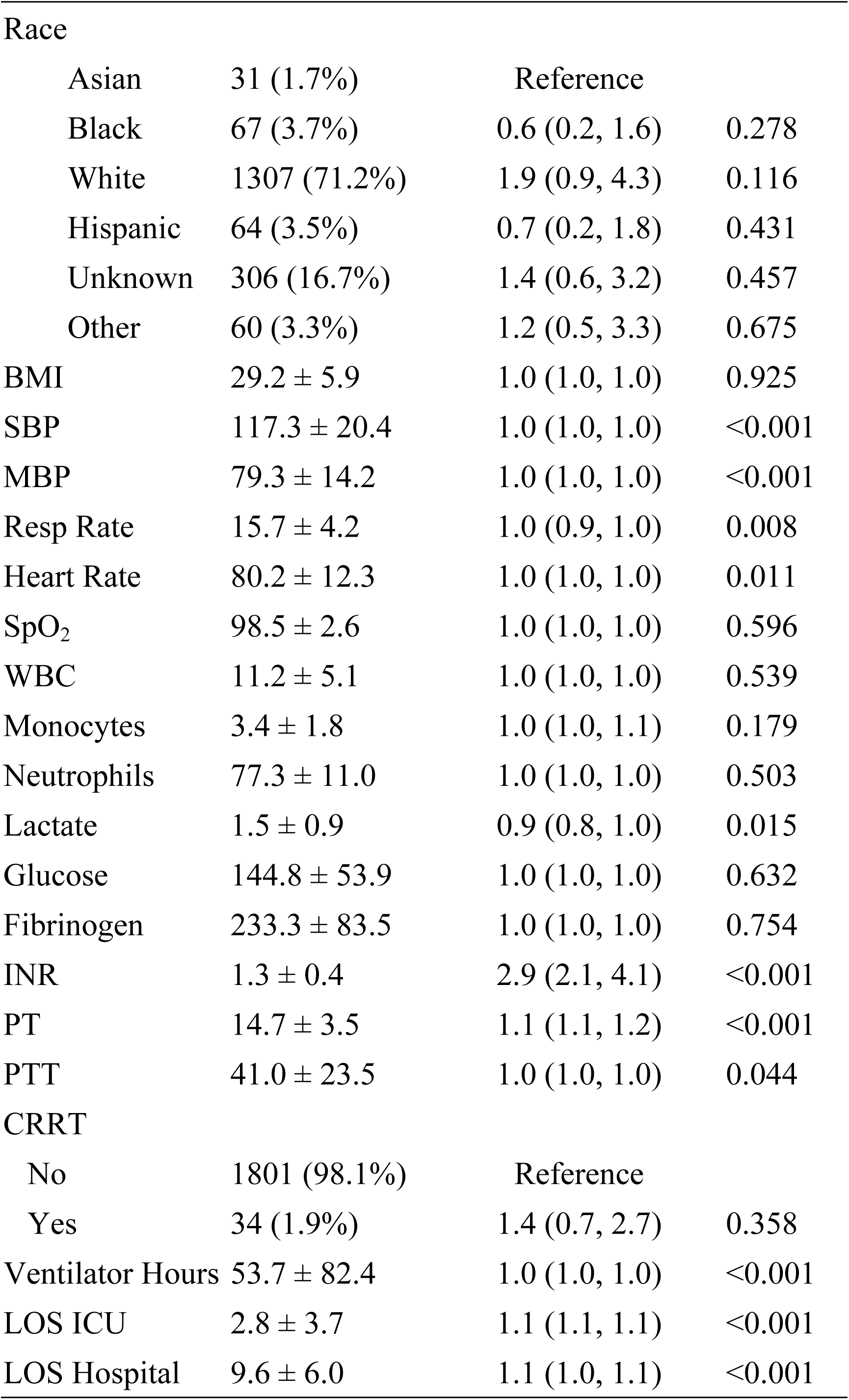
The results of univariate analysis.

| Variables | Statistics | AF (OR 95%CI) | P-Value |
| --- | --- | --- | --- |
| CABG | 0.8 ± 0.4 | 2.1 (1.6, 2.7) | <0.001 |
| Age | 69.5 ± 11.1 | 1.1 (1.1, 1.1) | <0.001 |
| Gender |  |  |  |
| Female | 1332 (72.6%) | Reference |  |
| Male | 503 (27.4%) | 1.0 (0.8, 1.2) | 0.994 |
| Race |  |  |  |
| Asian | 31 (1.7%) | Reference |  |
| Black | 67 (3.7%) | 0.6 (0.2, 1.6) | 0.278 |
| White | 1307 (71.2%) | 1.9 (0.9, 4.3) | 0.116 |
| Hispanic | 64 (3.5%) | 0.7 (0.2, 1.8) | 0.431 |
| Unknown | 306 (16.7%) | 1.4 (0.6, 3.2) | 0.457 |
| Other | 60 (3.3%) | 1.2 (0.5, 3.3) | 0.675 |
| BMI | 29.2 ± 5.9 | 1.0 (1.0, 1.0) | 0.925 |
| SBP | 117.3 ± 20.4 | 1.0 (1.0, 1.0) | <0.001 |
| MBP | 79.3 ± 14.2 | 1.0 (1.0, 1.0) | <0.001 |
| Resp Rate | 15.7 ± 4.2 | 1.0 (0.9, 1.0) | 0.008 |
| Heart Rate | 80.2 ± 12.3 | 1.0 (1.0, 1.0) | 0.011 |
| SpO<sub>2</sub> | 98.5 ± 2.6 | 1.0 (1.0, 1.0) | 0.596 |
| WBC | 11.2 ± 5.1 | 1.0 (1.0, 1.0) | 0.539 |
| Monocytes | 3.4 ± 1.8 | 1.0 (1.0, 1.1) | 0.179 |
| Neutrophils | 77.3 ± 11.0 | 1.0 (1.0, 1.0) | 0.503 |
| Lactate | 1.5 ± 0.9 | 0.9 (0.8, 1.0) | 0.015 |
| Glucose | 144.8 ± 53.9 | 1.0 (1.0, 1.0) | 0.632 |
| Fibrinogen | 233.3 ± 83.5 | 1.0 (1.0, 1.0) | 0.754 |
| INR | 1.3 ± 0.4 | 2.9 (2.1, 4.1) | <0.001 |
| PT | 14.7 ± 3.5 | 1.1 (1.1, 1.2) | <0.001 |
| PTT | 41.0 ± 23.5 | 1.0 (1.0, 1.0) | 0.044 |
| CRRT |  |  |  |
| No | 1801 (98.1%) | Reference |  |
| Yes | 34 (1.9%) | 1.4 (0.7, 2.7) | 0.358 |
| Ventilator Hours | 53.7 ± 82.4 | 1.0 (1.0, 1.0) | <0.001 |
| LOS ICU | 2.8 ± 3.7 | 1.1 (1.1, 1.1) | <0.001 |
| LOS Hospital | 9.6 ± 6.0 | 1.1 (1.0, 1.1) | <0.001 |

**Table 5.**
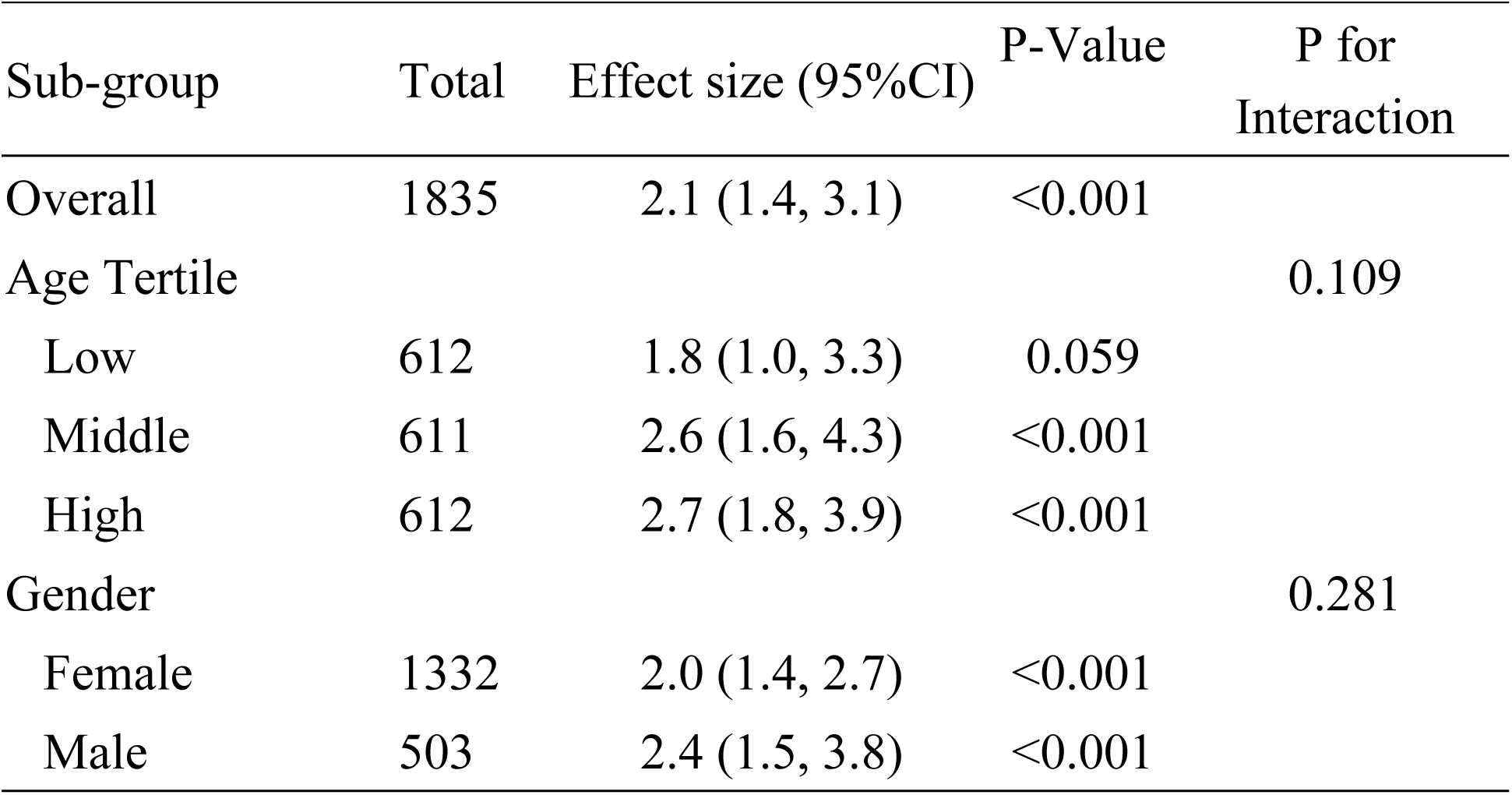
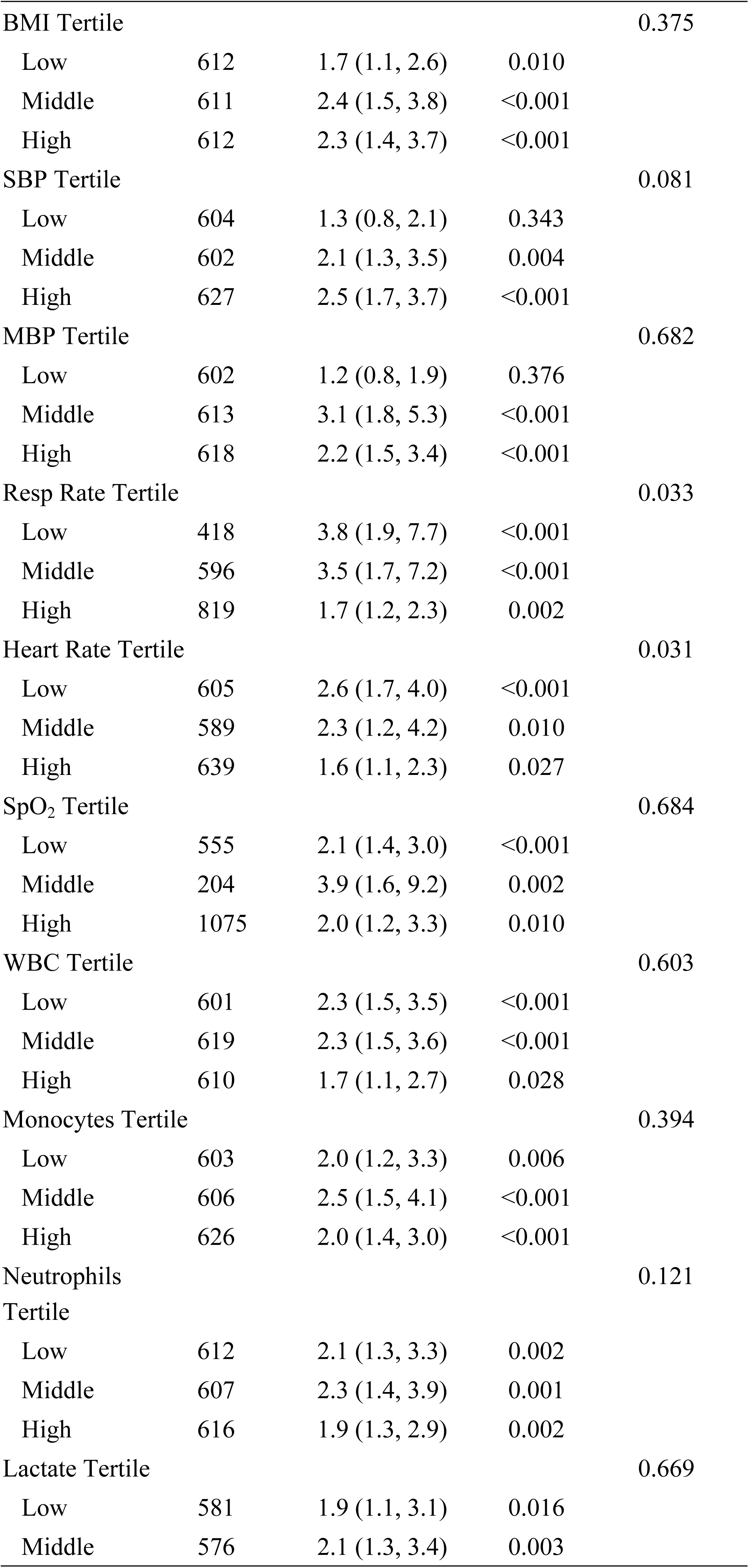
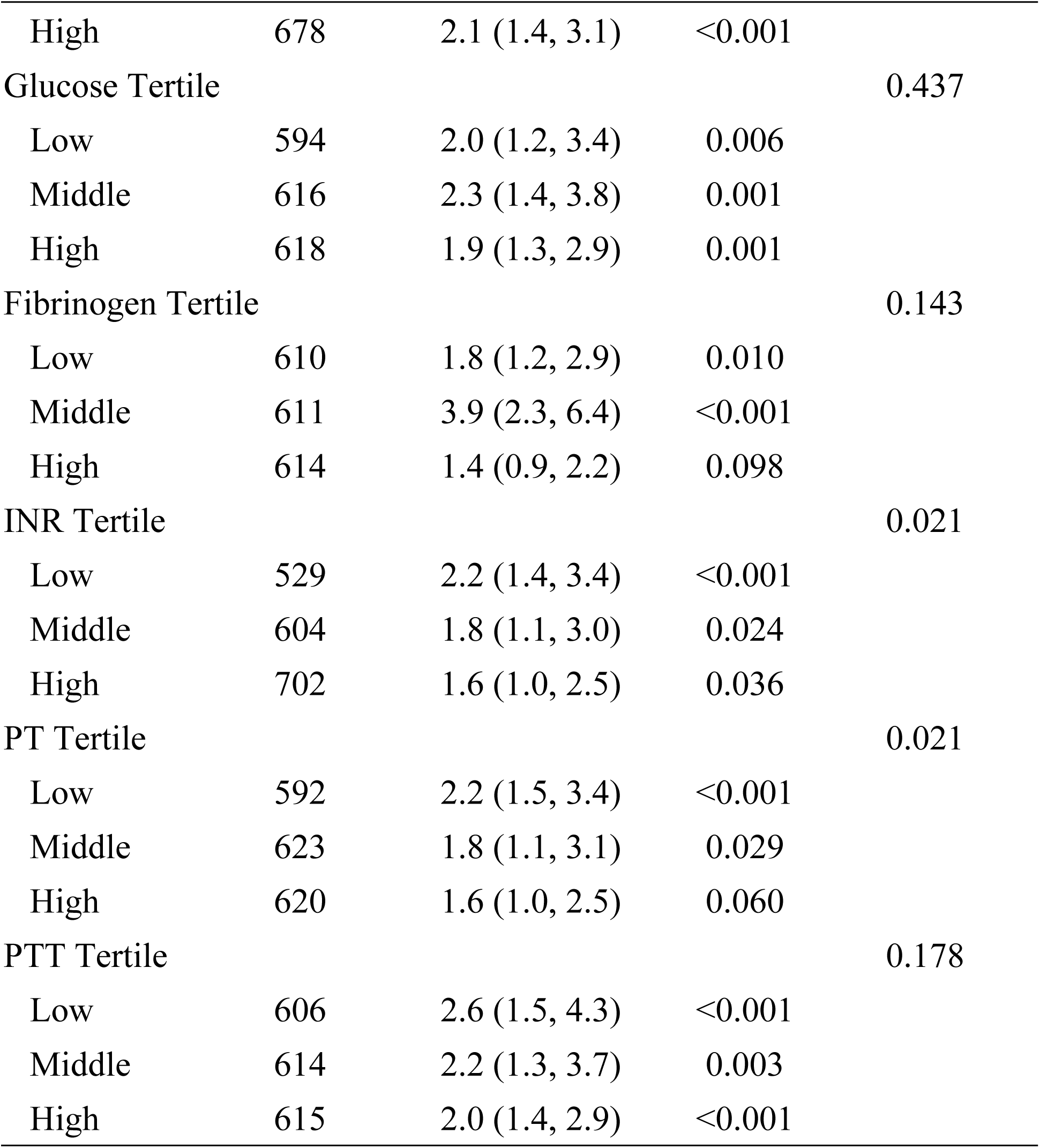
The results of subgroup analysis.

Respiratory rate and heart rate demonstrated significant interactions (P < 0.05), suggesting that these physiological factors may modify the association between CABG and AF. In particular, lower respiratory and heart rates were associated with a stronger effect of CABG on AF risk. Additionally, significant interactions were observed with INR and PT, indicating that coagulation status may influence the CABG–AF relationship.

The results of a multiple regression analysis examining the association between CABG and various clinical outcomes across three different models. Model I (Non-adjusted):OR for AF is 2.1 (95% CI: 1.6, 2.7), with a p-value <0.001, indicating a significant association between CABG and AF.Model II (Adjusted for age, gender, race, BMI): OR increases to 2.6 (95% CI: 1.9, 3.4), with a p-value <0.001, suggesting that after adjusting for basic demographic factors, the association remains significant.Model III (Fully adjusted): OR is 2.1 (95% CI: 1.4, 3.1), with a p-value <0.001, indicating that even after adjusting for a comprehensive set of clinical and laboratory variables, CABG is still significantly associated with AF(Table 6 shown).

**Table 6.**
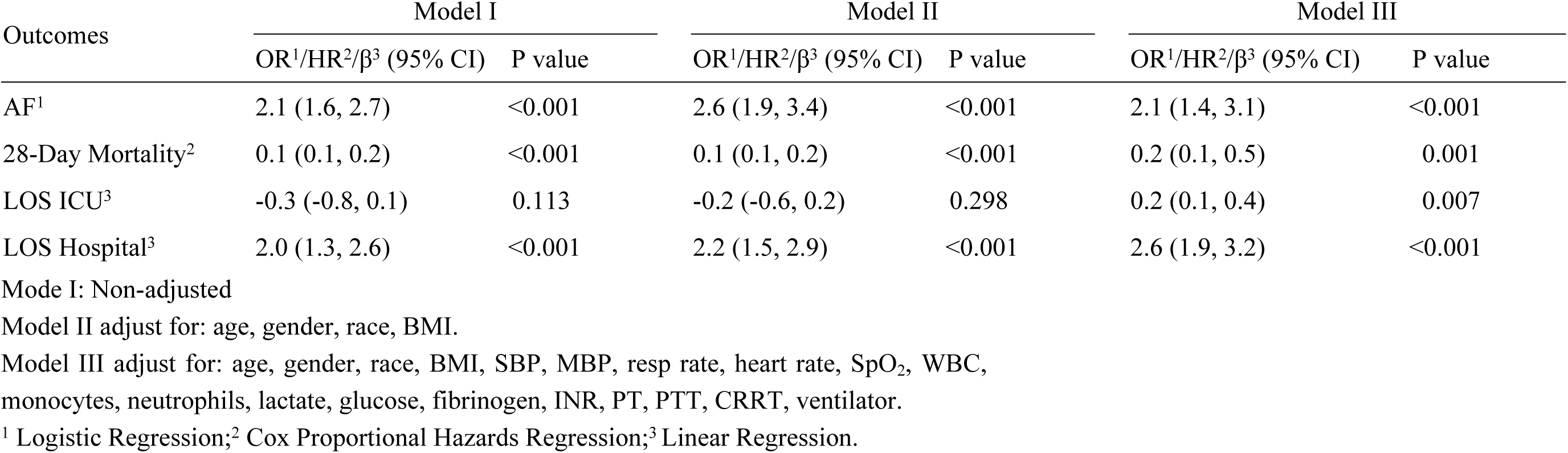
Multiple regression analysis of CABG and clinical outcomes.

## Discussion

While observational studies have identified associations between CABG and AF, confounding factors and reverse causality limit causal inference. Our study uniquely integrates MR and clinical cohort data to address this gap.

The MR analysis revealed a strong positive causal relationship between CABG and AF, with the IVW method yielding a highly significant p-value of 2.33 x 10⁻⁹. This suggests that CABG may increase the risk of AF, highlighting the importance of monitoring and managing AF in patients undergoing CABG. The consistency of results across multiple MR models (MR-Egger, weighted median) further strengthens the validity of our findings(Figure 2 shown).Multivariable MR (MVMR) analysis, which adjusted for confounders such as Type 2 diabetes, hypertension, pulmonary embolism, and hyperthyroidism, confirmed that the causal relationship between CABG and AF is independent of these factors(Table 2 shown). This causal link persisted even after adjusting for confounders such as diabetes and hypertension in multivariable MR, suggesting that CABG itself—rather than comorbidities—drives AF risk. These findings align with observational cohort results, where CABG patients exhibited over twice the risk of AF compared to non-CABG patients (OR = 2.1, 95% CI: 1.4–3.1, P < 0.001)(Table 4 shown), corroborating prior reports of post-CABG AF incidence (20–50%) and its association with adverse clinical outcomes (12–15).

Subgroup analyses revealed that the risk of AF tended to increase with higher age and systolic blood pressure, consistent with known risk factors for AF(Table 5 shown). Interestingly, lower respiratory and heart rates were associated with a stronger effect of CABG on AF risk, suggesting that autonomic nervous system modulation may play a role in the development of post-operative AF. Additionally, interactions with INR and PT indicated that coagulation status may influence the CABG-AF relationship, highlighting the complex interplay between surgical stress, inflammation, and hemostasis in the pathogenesis of AF(4, 16, 17).

AF is a serious complication following CABG, associated with adverse outcomes such as stroke, heart failure, increased mortality, prolonged hospital stays, and higher healthcare costs(2, 18). The EXCEL trial had reported a higher incidence of AF after CABG compared to percutaneous coronary intervention (PCI) in patients with left main coronary artery disease (19). Additionally, research by Professors Alatassi demonstrated that while AF did not increase perioperative mortality, active ablation of AF could improve mid- and long-term survival (20). The cohort studies, along with our results, emphasize the importance of addressing AF as a critical complication of CABG.

The identification of CABG as an independent risk factor for AF has important implications for clinical practice. It underscores the need for vigilant monitoring of AF in patients undergoing CABG and highlights the potential benefits of prophylactic strategies to reduce the incidence of AF(21–23).

Our study has several methodological strengths. The cohort study results are consistent with the findings from the MR analysis, which provided robust evidence of a causal relationship between CABG and AF. The convergence of results from both approaches strengthens the validity of our conclusions and highlights the importance of CABG as a risk factor for AF. Therefore, while the cohort study demonstrated an association between CABG and AF, it cannot establish causality. The MR analysis complements this limitation by providing evidence of a causal relationship.

Despite these insights, our study has limitations. First, the European ancestry of GWAS data limits generalizability to other populations, necessitating validation in diverse cohorts. While MR supports causality, the retrospective cohort design risks residual confounding. Second, the MR analysis assumes linearity of genetic effects, which may not capture non-CABG-related triggers (e.g., emergency surgery).Third, the MIMIC-IV database’s ICU focus may restrict applicability to non-critically ill populations and the MIMIC-IV cohort primarily includes ICU patients, potentially overestimating AF risk compared to general surgical populations. Finally, the precise biological mechanisms linking CABG to AF remain speculative. Although we hypothesize roles for inflammation, oxidative stress, and ANS modulation, direct evidence is lacking.

Our findings advocate for proactive AF surveillance in CABG patients, particularly those with advanced age, hypertension, or autonomic instability. Perioperative interventions targeting inflammation or ANS modulation warrant exploration to mitigate AF risk. Furthermore, the development of risk prediction models incorporating genetic and clinical factors could help identify patients at high risk of AF and guide personalized prevention strategies. Finally, expanding MR analyses to include diverse populations and individual-level data could enhance the generalizability and clinical applicability of these findings.

### Conclusion

The cohort study provides compelling evidence of a significant association between CABG and the development of AF, particularly NOAF. By combining MR and clinical cohort analyses, this study robustly establishes CABG as an independent risk factor for NOAF. Future research should validate these findings in multi-ethnic cohorts and explore targeted interventions (e.g., anti-inflammatory therapies) to mitigate AF risk

## Consent for publication

All authors consent to publication of this article.

## Data Availability

All relevant data are within the manuscript and its Supporting Information files.

